# Determinants of early initiation and exclusive breastfeeding of the Bhumij community of India: a community-based cross-sectional study

**DOI:** 10.64898/2026.07.15.26358159

**Authors:** Debasree Das, Chandana Basu Mallick, Brijesh P. Singh, Arup Ratan Bandyopadhyay

## Abstract

**Background:** Breastfeeding practices vary across populations and are influenced by a range of social-cultural, and healthcare-related factors. Evidence on early initiation of breastfeeding (EIBF) and exclusive breastfeeding (EBF) among indigenous communities in India remains limited. This study aimed to identify factors associated with EIBF and EBF among Bhumij mothers in eastern India.

**Method:** A community-based cross-sectional study of 306 Bhumij mother-child pairs was conducted in Purulia, West Bengal, India (2023-2024). Socio-demographic and breastfeeding-related data were collected through structured interviews and analysed using bivariate and multivariable logistic regression.

**Results:** Almost all children (99.3%) had been breastfed; 72.9% initiated breastfeeding within one hour of birth, and 65.2% were exclusively breastfed during the first six months. In multivariable analyses, mode of delivery, pre-lacteal feeding, and colostrum discarding were significantly associated with EIBF. Maternal knowledge of EBF was positively associated with EBF practice (adjusted OR: 6.14; 95% CI: 2.80-13.46). Maternal perception of milk production was also associated with EBF, with higher odds observed among mothers reporting adequate (adjusted OR: 6.12; 95% CI: 3.00-12.48) or profuse (adjusted OR: 7.71; 95% CI: 2.56-23.25) milk production compared with those reporting insufficient milk production.

**Conclusion:** This study provides evidence on breastfeeding practices among Bhumij mothers and identifies healthcare-related, maternal, and caregiving factors associated with EIBF and EBF. The findings contribute to the limited literature on infant feeding practices among indigenous populations in India and may inform breastfeeding promotion and nation-wide maternal-child health programmes in similar settings.

## Introduction

Breastfeeding provides a natural source of nutrition that supports an infant’s growth, development, maturation, and survival [1]. Studies demonstrated that it significantly reduces child mortality and prevents morbidities and, in the long term may decrease obesity and diabetes in later life [2–5]. It also provides substantial maternal benefits by reducing postpartum complications, and long-term health risks, and supporting postpartum recovery [1,6]. The World Health Organization (WHO) and the United Nations Children’s Fund (UNICEF) recommend that breastfeeding begin immediately after birth, often referred to as early initiation of breastfeeding (EIBF), implying that infants are offered the breastmilk within the first hour after delivery. This further facilitates the timely provision of colostrum- the initial thick, yellowish milk, rich in essential nutrients and immunoglobulins, serving as the infant’s first source of nutrition and immunity. The WHO recommends exclusive breastfeeding (EBF), which is defined as feeding infants only breast milk of the mother or a wet nurse for the first six months, without additional food or liquids except the oral rehydration solution or drops/syrups of vitamins, minerals, or medicines. After six months, appropriate complementary foods should be introduced while continuing breastfeeding up to two years or beyond [7].

Globally, only 48% of infants were exclusively breastfed in 2023 (UNICEF, 2023) [8]. Breastfeeding practices have cross cultural variations such as, considerably across low- and middle-income countries (LMICs) [9–11]. Neonatal mortality rates also continue to differ across regions, with estimates of 26 and 21 per 1,000 live births in Sub-Saharan Africa and Southern Asia, respectively (UNIGME, 2024). Breastfeeding is widely recognized as a cost-effective and evidence-based intervention that supports infant health and survival by reducing early-life morbidity and mortality [12].

Traditional beliefs and customary infant feeding practices have been reported across societies for generations [13]. Several contextual and healthcare-related factors may delay the early initiation of breastfeeding in low- and middle-income countries (LMICs) [14–18]. Cultural beliefs and customary practices regarding colostrum may also influence the timing of breastfeeding initiation, which can affect early infant feeding patterns and newborn health outcomes [19,20]. Additionally, many infants receive liquids other than breast milk during the first three days after birth [21–24]. In countries such as India, Bangladesh, and Nepal, prelacteal feeding practices have been reported at rates of 77%, 49.9%, and 26.5%, respectively [25].

Approximately 8.6% of India’s population belongs to the Scheduled Tribes (STs), representing socio-culturally diverse communities with varying health and healthcare experiences (Census of India, 2011). Differences in access to education, maternal healthcare services, and health-related information may influence neonatal and infant health outcomes across these populations. In addition, community traditions, cultural values, and locally practiced caregiving approaches play an important role in shaping breastfeeding behaviours and maternal-child health practices. West Bengal has an estimated under-five mortality rate of 25.4 per 1,000 live births, showing improvement from earlier NFHS-5 estimates (IIPS, 2020).

Bhumij, one of the major indigenous ethnic groups in India, is primarily concentrated in the states of Jharkhand, West Bengal, and Odisha. Variations in access to reproductive healthcare services, health-related information, and breastfeeding support may influence child nutrition and feeding outcomes among preschool children in the Bhumij community [26]. The Bhumij community also follows socio-cultural traditions that shape infant-feeding practices, including the ritual offering of first milk (colostrum) to the earth, reflecting origin narratives and cosmological beliefs associated with the soil. Given the established importance of exclusive breastfeeding (EBF) for child survival, particularly in resource-constrained settings, this study provides the first comprehensive assessment of breastfeeding practices among the Bhumij community in the Purulia district.

## Methodology

### Study Design

A community-based cross-sectional study was conducted among the Bhumij community in the Baghmundi Block of Purulia District, West Bengal, India. The Bhumij, a recognised Scheduled Tribe (ST), have a high population concentration in this area; therefore, the block was purposely selected to ensure adequate representation. Data were collected from more than 32 villages selected through random sampling.

### Study Population

The study population comprised women of childbearing age, 15 to 45 years, who had a living child aged ≤30 months at the time of data collection. This criterion represents the reproductive age group and early infancy period when breastfeeding behaviours, particularly EIBF and EBF, are most critical. Eligible participants were identified through household visits, and a random sampling approach was used to select households within each village.

### Data Collection

Data were collected through a semi-structured interview guide, with assistance from frontline health workers at different primary health centres (PHCs) in Baghmundi block, between March 2023 and April 2024. Information on socio-demographic characteristics, infant feeding practices, and maternal reproductive and obstetric-related factors was obtained. All interviews were conducted in the local language.

### Ethical Considerations

Written informed consent was obtained from all participants prior to data collection. For pre-literate respondents, consent was recorded *via* left thumb impression in the presence of an independent witness. Ethical approval for this study was obtained from the Institutional Ethical Committee for Biomedical and Health Research involving Human Participants, University of Calcutta (Ethical Clearance No. CUIEC/02/16/2022-23).

### Sample size estimation

In the present study, the dependent variable was EBF status among Bhumij mothers. Due to the unavailability of published data for the Bhumij community, sample participants size estimation was based on the rural West Bengal EBF prevalence of 65.1% reported in NFHS-5 (2019–2021). Based on this proportion, a minimum sample size of 384 was estimated for a 95% confidence level and a 0.05 margin of error. However, owing to field constraints, data were collected from 306 Bhumij mothers, yielding which corresponds to an achievable margin of error of approximately 5.6%. Recruitment was facilitated through community health workers and outreach teams from one Block Primary Health Centre (BPHC) and three Primary Health Centres (PHCs) including Accredited Social Health Activists (ASHA) and Auxiliary Nurse Midwives (ANM) within the Baghmundi block.

### Study variables

Data were collected through household visits using a semi-structured, pre-tested schedule comprising both closed- and open-ended questions. Interviews were conducted in the local language, thereby, ensuring privacy and accuracy. Mothers who were critically ill were excluded. The study aimed to assess breastfeeding practices and factors affecting them, including socio-demographic, maternal obstetric and childbirth-related, and infant feeding behaviour-related determinants.

The study had two principal dependent variables, such as EIBF within one hour and EBF up to six months. The variables were broadly categorised into four categories viz demographic and socio-economic factors, maternal reproductive and pre-childbirth characteristics, child-birth related characteristics and breastfeeding-related characteristics. EIBF was defined as putting the newborn to the breast immediately after birth. Mothers were asked, “How long after birth did you first put your baby to the breast?”, and the time was recorded in hours or days following standard field instructions. Initiation was coded as “early” when breastfeeding began within one hour of birth and as “delayed” when initiation occurred after the first hour. EBF was defined as feeding the infant only breastmilk for the first six months of life, and mothers were asked whether their child had been exclusively breastfed up to six months; responses were coded as “yes” or “no”. Explanatory variables included maternal socio-demographic factors (mother’s age, education, employment status, monthly family expenditure, and anthropometric measurements of height (cm) and weight (kg) using standard protocol to derive BMI) [27]; maternal reproductive and pre-childbirth characteristics (age at menarche, age at first pregnancy, parity; “primiparous” or “multiparous”, antenatal care (ANC) visits <4 or ≥4, antenatal morbidities were coded to indicate whether the outcome was present or not (“Yes” or “No”), whether having any history of breast diseases during index child’s pregnancy, and supplements intake during pregnancy; childbirth-related characteristics include labour experience, place of delivery, type of delivery; “assisted institutional vaginal birth”, “Caesarean birth” or “unassisted vaginal birth/home delivery”, gestational age, type of pregnancy, and birth order; and child characteristics incorporated newborn morbidity, sex of infant’s sex, birth weight, and current age. Breastfeeding-related explanatory variables assessed whether mothers received counselling on breastfeeding before starting breastfeeding, their primary information source, antenatal preparation for breastfeeding, colostrum feeding behaviour; “discarded” or “not-discarded”, reasons for colostrum discard; “elder’s advice”, “bad for health”, baby can’t suck”, prelacteal feeding practices (liquid other than the breastmilk), the first feed offered after birth, maternal knowledge regarding EBF, breastfeeding frequency in a 24-hour period, breastfeeding on demand, the usual time of day when the infant fed for the longest duration; “ Day” or “Night”, and the mother’s self-perception of breastmilk production.

### Statistical analysis

Data were coded and analysed using the SPSS package version 27.0 (SPSS Inc., Chicago, IL, USA). Data are presented as means with SD or as counts (n) with percentages. The odds ratios (ORs) with 95% confidence intervals were estimated using binary logistic regression to assess the association between each independent variable and the dependent variable. The results of the bivariate analysis were also used to guide variable selection for the multivariable logistic regression model. Following standard epidemiological practice, variables with p-values < 0.30 in the bivariate analysis were included in the multivariable model, and those exceeding this threshold were excluded. Both binary and multivariable logistic regression analyses were conducted to evaluate the likelihood of EIBF and EBF across categories of explanatory variables. Adjusted Odds Ratios (AORs) with 95% confidence intervals were generated in the multivariable model to estimate the independent effect of each predictor.

## Results

### Demographic and socio-economic characteristics of the study participants and infants

The study included 306 mother-child pairs, with a response rate of 100%. Mothers had a mean age of 23.72 ± 3.23 years (range: 13–37). Most families (76.47%) reported a monthly household income below 5000. A large proportion of mothers were dedicated to household responsibilities (94.8%), and over half (52.3%) had attained secondary-level education. The mean age of the infants was 11.65 ± 3.26 months, and 32.8% had a birth weight below 2500 g. Male infants constituted 54% of the study population, while only 15.7% had a history of neonatal morbidity (Table 1).

**Table 1:** Socio-demographic characteristics of the study participants and infants.

| Demographic and socio-economic characteristics | Mean(SD)*n(%)# | Infant's characteristics | Mean(SD)*n(%)# |
| --- | --- | --- | --- |
| Mother's age* | 23.72(3.23) | Newborn morbidity# |  |
| Educational qualification# |  | No | 258(84.31) |
| Pre-literate | 65(21.24) | Yes | 48(15.69) |
| Primary | 33(10.8) | Sex of infant# |  |
| Secondary | 160(52.3) | Male | 166(54.25) |
| Higher studies | 48(15.7) | Female | 140(45.75) |
| Mother's employment status# |  | Child's Birth weight* | 2545.4(445.6) |
| Working | 16(5.23) | Newborn birtweight# |  |
| Non-working | 290(94.77) | <2500grms | 98 (32.8) |
| Per-month family expenditure# |  | >2500grms | 201 (67.2) |
| <5000 | 234(76.47) | Child's age* | 11.65(3.26) |
| ≥5000 | 72(23.53) | **Breastfeeding-related characteristics** |  |
| Mother's height* | 145.08(5.23) | Counseling on BF# |  |
| Mother's weight* | 42.18(5.8) | Yes | 241(78.75) |
| Mother's BMI# |  | No | 65(21.25) |
| Normal weight | 221(72.22) | Source of information# |  |
| Underweight | 68(22.22) | Health workers | 185(60.46) |
| Overweight | 9(2.94) | Family-neighbours | 66(21.57) |
| Obese | 8(2.61) | None | 55(17.97) |
| **Mother's reproductive and pre-childbirth characteristics** |  | Antenatal preparation for BF# |  |
| Age at menarche* | 12.42(1.3) | Yes | 158(51.63) |
| Age at first pregnancy* | 17.96(2.1) | No | 148(48.36) |
| Parity# |  | EIBF# |  |
| Primiparous | 125(59.15) | <1 hour | 223(72.9) |
| Multiparous | 181(40.85) | >1 hour | 83(27.12) |
| ANC visit# |  | Colostrum discarded# |  |
| ≥4 | 236(77.12) | Yes | 208(67.97) |
| <4 | 70(22.9) | No | 98(32.03) |
| Antenatal morbidities# |  | Reason for discarding colostrum# |  |
| Yes | 244(79.74) | Bad for health | 83(27.12) |
| No | 62(20.26) | Elder's advice | 100(32.7) |
| Any breast disease# |  | Baby can't suck | 25(8.17) |
| Yes | 4(1.31) | None | 98(32.02) |
| No | 302(98.69) | Prelacteal feed# |  |
| Supplements during pregnancy# |  | No | 270(88.24) |
| Yes | 289(94.44) | Yes | 36(11.76) |
| No | 17(5.56) | First fed with# |  |
| Labour experience# |  | Water | 7(2.29) |
| Normal labour | 232(75.82) | Formula milk | 25(8.17) |
| Short labour | 62(20.3) | Animal milk | 4(1.31) |
| Prolonged labour | 12(3.92) | Colostrum | 93(30.4) |
| Place of delivery# |  | Breastmilk | 177(57.84) |
| Govt | 267(87.25) | Knowledge of EBF# |  |
| Home | 27(8.82) | Yes | 234(76.47) |
| Private | 12(3.92) | No | 72(23.53) |
| Type of delivery# |  | EBF upto 6m# (n=244) |  |
| AIVB | 250(81.7) | Yes | 159(65.16) |
| C/S | 29(9.48) | No | 85(34.83) |
| UVB | 27(8.82) | BF frequency per 24hour# |  |
| Gestational age# |  | <8 | 79(25.82) |
| Term | 249(81.37) | 8-12 | 152(49.67) |
| Pre-Term | 57(18.63) | >12 | 71(23.2) |
| Type of pregnancy# |  | NA | 4(1.31) |
| Singleton | 296(96.73) | Time to BF# |  |
| Multiple | 10(3.27) | NA | 4(1.31) |
| Birth order# |  | Baby cries | 242(79.08) |
| 1 | 123(40.2) | Mother's convenience | 60(19.61) |
| 2-3 | 152(49.67) | Longest time baby fed# |  |
| >3 | 31(10.13) | Day | 215(70.26) |
|  |  | Night | 87(28.43) |
|  |  | NA | 4(1.31) |
|  |  | Self-perception of Breastmilk production# |  |
|  |  | Inadequate | 104(33.98) |
|  |  | Adequate | 151(49.35) |
|  |  | Profuse | 51(16.66) |

\*- Mean(SD); #- Frequency(%)
BMI- Body Mass Index
ANC-ante Natal Care
AIVB- Assisted institutional Vaginal Birth
C/S- Cesarean section
UVB- Unassisted Vaginal Birth
BF- Breastfeeding
EIBF- Early Initiation of Breastfeeding
EBF-Exclusive Breastfeeding
NA- Not applicable

### Mother’s reproductive and pre-childbirth characteristics

The obstetric profile of the 306 study participants indicated a mean age at menarche of 12.4 ± 1.3 years and a mean age at first pregnancy of 17.96 ± 2.1 years. More than half of the mothers (59.15%) were primiparous, and 77.12% reported attending four or more antenatal care (ANC) visits during their most recent pregnancy. The majority of deliveries occurred in institutional health facilities (91.18%) and were vaginal (90.52%). Nutritional supplementation during pregnancy was reported by 94.4% of mothers (Table 1).

### Breastfeeding practices

Almost all children (n = 304, 99.34%) had been breastfed at some point during infancy. Breastfeeding was initiated within one hour of birth for 72.9% of infants. Postpartum breastfeeding counselling from trained healthcare workers (ASHAs) was reported by 78.75% of mothers. Healthcare workers were the most frequently reported source of breastfeeding information (60.46%), followed by family members and neighbours (21.6%). (Table 1).

Among the participants, 234 (76.5%) mothers had knowledge regarding exclusive breastfeeding (EBF) recommendations. 65.16% (n = 159) reported practicing EBF for the recommended first six months, indicating variation between breastfeeding knowledge and reported feeding practices. For the assessment of EBF, children aged 6 months and above were considered in accordance with the UNICEF/WHO recommendations for the first six months. Of the 244 participants, 65.16% of children were exclusively breastfed at the time of survey.

Colostrum discarding was reported by 67.9% of participants. Commonly reported reasons included perceived infant health concerns, advice from family elders, and difficulties with infant suckling. These findings suggest that neonatal feeding practices may be influenced by a combination of family, cultural, and caregiving factors. In contrast, pre-lacteal feeding was reported by 11.8% of participants (n = 36).

### Factors affecting early initiation of breastfeeding (EIBF)

Retrospective assessment indicated that almost all participating mothers (99.34%, n = 304) had initiated breastfeeding. Early initiation of breastfeeding (EIBF), defined as initiation within the first hour after birth, was reported by 72.9% (n = 223) of mothers. A further 12.1% (n = 37) initiated breastfeeding within 2–3 hours after delivery, while 3.9% (n = 12) reported initiation later on the day of birth. Breastfeeding initiation after the first day of life was reported by 10.4% (n = 32) of participants (Table 2).

**Table 2:** Time to initiation of breastfeeding after birth (%)

| Time to start breastfeeding | Frequency (n) | Percentage |
| --- | --- | --- |
| <1hr | 223 | 72.9 |
| 2-3hrs | 37 | 12.1 |
| <24hrs | 12 | 3.9 |
| >1day | 32 | 10.4 |

Analysis of breastfeeding initiation timing demonstrated a significant association with EIBF practices (p < 0.001). Factors associated with EIBF were further examined using bivariate and multivariable logistic regression analyses (Table 3). In the multivariable logistic regression analysis, mode of delivery emerged as the strongest determinant of early initiation of breastfeeding. Mothers who delivered by caesarean section had lower odds of initiating breastfeeding within the first hour after birth compared with those who had a vaginal delivery (adjusted OR: 0.12; 95% CI: 0.05–0.30), corresponding to an approximately 88% reduction in the odds of early initiation of breastfeeding.

**Table 3:** Determinants of early initiation of breastfeeding (EIBF) among Bhumij mothers. Bivariate and multivariable logistic regression model predicting the likelihood of practicing early initiation of breastfeeding

| Variable | OR(95% CI) | p-value | AOR(95%CI) | p-value |
| --- | --- | --- | --- | --- |
| *Educational qualification* |  |  |  |  |
| Literate (241) | 1.424(0.784-2.585) | 0.246 | 1.285(0.601-2.746) | 0.517 |
| Pre-literate (65) | Reference |  |  |  |
| *Occupation* |  |  |  |  |
| Working (16) | 0.445(0.160-1.236) | 0.12 | 0.4(0.115-1.388) | 0.149 |
| Non-working (290) | Reference |  |  |  |
| *Monthly income* |  |  |  |  |
| ≥5000(72) | 0.904(0.499-1.638) | 0.74 |  |  |
| <5000 (234) | Reference |  |  |  |
| *Parity* |  |  |  |  |
| Multiparous (181) | 1.146(0.685-1.917) | 0.605 |  |  |
| Primiparous (125) | Reference |  |  |  |
| *ANC visits* |  |  |  |  |
| ≥ 4 visits (236) | 0.736(0.410-1.320) | 0.303 | 0.973(0.465-2.038) | 0.943 |
| <4 (70) | Reference |  |  |  |
| *Ante Natal Morbidity (ANM)* |  |  |  |  |
| Yes (244) | 1.413(0.770-2.591) | 0.264 |  |  |
| No (62) | Reference |  |  |  |
| *Labour experience* |  |  |  |  |
| Normal (232) | 0.74(0.382-1.434) | 0.373 |  |  |
| Prolonged (12) | 1.458(0.286-7.449) | 0.65 |  |  |
| Short (62) | Reference |  |  |  |
| *Place of birth* |  |  |  |  |
| Institutional (279) | 1.177(0.494-2.804) | 0.713 |  |  |
| Non-institutional (27) | Reference |  |  |  |
| *Type of delivery* |  |  |  |  |
| C Section (29) | 0.093(0.038-0.228) | <0.0018** | 0.133(0.048-0.369) | <.001*** |
| Vaginal (277) | Reference |  |  |  |
| *Newborn morbidity* |  |  |  |  |
| Yes (258) | 0.197(0.103-0.378) | <0.001*** | 0.447(0.194-1.031) | 0.059 |
| No (48) | Reference |  |  |  |
| *Gestational Age* |  |  |  |  |
| Pre-term (57) | 0.283(0.155-0.516) | <.001*** | 0.476(0.218-1.041) | 0.063 |
| Term (249) | Reference |  |  |  |
| *Newborn birthweight* |  |  |  |  |
| >2500grms (201) | 1.298(0.760-2.214) | 0.339 |  |  |
| <2500grms (98) | Reference |  |  |  |
| *Newborn's sex* |  |  |  |  |
| Male (166) | 0.949(0.569-1.583) | 0.841 |  |  |
| Female (140) | Reference |  |  |  |
| *Mother's BMI* |  |  |  |  |
| Normal (221) | 0.947(0.506-1.772) | 0.866 |  |  |
| Overweight (8) | 0.417(0.100-1.732) | 0.228 |  |  |
| Obese (9) | 0.556(0.120-2.573) | 0.452 |  |  |
| Underweight (68) | Reference |  |  |  |
| *Counseling on breastfeeding* |  |  |  |  |
| Yes (241) | 1.385(0.784-2.446) | 0.262 | 0.584(0.287-1.189) | 0.138 |
| No (65) | Reference |  |  |  |
| *Colostrum discard* |  |  |  |  |
| Yes (208) | 0.346(0.183-0.653) | 0.001*** | 0.333(0.152-0.730) | 0.006* |
| No (98) | Reference |  |  |  |
| *Prelacteal feeding* |  |  |  |  |
| Yes (36) | 0.094(0.042-0.213) | <.001 | 0.146(0.059-0.364) | <.001*** |
| No (270) | Reference |  |  |  |
| *Self-perception on BM production* |  |  |  |  |
| Adequate (151) | 2.035(1.164-3.558) | 0.013** | 1.503(0.742-3.043) | 0.258 |
| Profuse (51) | 2.07(0.949-4.515) | 0.067 | 1.403(0.522-3.768) | 0.502 |
| Inadequate (104) | Reference |  |  |  |
| *0.05; **0.01; ***0.001 |  |  |  |  |

Note: The multivariable logistic regression model was adjusted for Educational qualification, Occupation, ANC visits, Type of Delivery, Neonatal morbidity, Gestational age, Counseling on breastfeeding, Colostrum discard, prelacteal feeding and Self-perception on BM production

Immediate postnatal feeding practices also showed a significant influence on breastfeeding initiation. Mothers who provided prelacteal feeds were substantially less likely to initiate breastfeeding early (OR: 0.38; 95% CI: 0.19–0.76), while mothers who discarded colostrum had nearly 50% lower odds of timely initiation (OR: 0.48; 95% CI: 0.24–0.96). Interestingly, exposure to breastfeeding information and antenatal counselling did not demonstrate a significant association with early initiation, suggesting that the counselling provided by healthcare workers alone may be insufficient to combat immediate delivery-related barriers and long-standing cultural practices. Collectively, the findings underscore that both delivery mode and early newborn feeding customs are critical determinants of breastfeeding initiation, emphasizing the need for strengthened post-caesarean lactation support (Table 3).

In the bivariate logistic regression analysis, several maternal, newborn, and health service-related factors were associated with early initiation of breastfeeding (EIBF). However, after adjustment for potential confounders in the multivariable model, only a subset of these factors remained significantly associated with EIBF (Table 3).

### Factors affecting exclusive breastfeeding (EBF)

In the multivariable logistic regression analysis, maternal knowledge of exclusive breastfeeding (EBF) was significantly associated with EBF practice. Mothers with adequate knowledge of EBF had higher odds of exclusively breastfeeding their infants compared with those with lower levels of EBF-related knowledge (adjusted OR: 6.14; 95% CI: 2.80–13.46; p < 0.001) (Table 4).

**Table 4:** Determinants of exclusive breastfeeding (EBF) among Bhumij mothers. Bivariate and multivariable logistic regression model predicting the likelihood of practicing exclusive breastfeeding

| Variable | OR(95%CI) | p-value | AOR(95%CI) | p-value |
| --- | --- | --- | --- | --- |
| *Maternal age (years)* | 0.978(0.918 -1.043) |  |  |  |
| *Educational qualification* |  |  |  |  |
| Literate (241) | 1.189 |  |  |  |
| Pre-literate (65) | Reference |  |  |  |
| *Occupation* |  |  |  |  |
| Working (16) | 0.516(0.161-1.653) | 0.266 | 2.38(0.42 - 13.85) | 0.251 |
| Non-working (290) | Reference |  |  |  |
| *Maternal morbidity* |  |  |  |  |
| Yes (244) | 0.892(0.481 - 1.653) | 0.717 |  |  |
| No (62) | Reference |  |  |  |
| *Parity* |  |  |  |  |
| Multiparous (181) | 0.761(0.444 - 1.303) | 0.319 |  |  |
| Primiparous (125) | Reference |  |  |  |
| *ANC visits* |  |  |  |  |
| ≥ 4 visits (236) | 1.162(0.626 -2.16) | 0.634 |  |  |
| <4 (70) | Reference |  |  |  |
| *Place of birth* |  |  |  |  |
| Institutional (279) | 1.548(0.587 - 4.082) | 0.377 |  |  |
| Non-institutional (27) | Reference |  |  |  |
| *Type of delivery* |  |  |  |  |
| C Section (29) | 0.503(0.201 -1.262) | 0.143 | 0.752(0.219 -2.578) | 0.752 |
| Vaginal (277) | Reference |  |  |  |
| *Newborn's sex* |  |  |  |  |
| Male (166) | 0.733(0.432 - 1.243) | 0.249 | 0.843(0.425 -1.675) | 0.626 |
| Female (140) | Reference |  |  |  |
| *Newborn birthweight* | 1.001(1.0 -1.001) | 0.069 | 1(1.0 -1.001) | 0.317 |
| *Knowledge of EBF* |  |  |  |  |
| Yes (234) | 6.038(3.147 -11.584) | 0.01** | 6.136(2.796 - 13.464) | 0.001*** |
| No (72) | Reference |  |  |  |
| *Self-perception on BM production* |  |  |  |  |
| Adequate (151) | 6.547(3.522-12.172) | <0.01** | 6.12(3.002 - 12.476) | 0.01** |
| Profuse (51) | 8.437(3.168-22.471) | <0.01** | 7.709(2.556 - 23.248) | 0.01** |
| Inadequate (104) | Reference |  |  |  |
| *Maternal BMI* |  |  |  |  |
| Normal (221) | 0.761(0.387 – 1.496) | 0.428 | 0.762(0.327 – 1.777) | 0.53 |
| Overweight (8) | 0.25(0.053 – 1.181) | 0.08 | 0.194(0.028 – 1.357) | 0.099 |
| Obese (9) | 1.042(0.182 – 5.976) | 0.963 | 0.481(0.064 – 3.642) | 0.479 |
| Underweight (68) | Reference |  |  |  |

\*0.05; \*\*0.01; \*\*\*0.001
Note: Multivariable logistic regression model was adjusted for Occupation, Type of delivery, Newborn sex, Child's birthweight, Knowledge of breastfeeding, Self-perception on BM production.

One of the interesting findings was that the maternal perception of breastmilk production was significantly associated with exclusive breastfeeding (EBF) practices. Mothers who perceived their milk secretion as adequate had over six-fold higher odds of practicing EBF (AOR: 6.12; 95% CI: 3.00–12.48), while those reporting profuse milk secretion demonstrated the highest likelihood of EBF, with nearly eight-fold increased odds (AOR: 7.71; 95% CI: 2.56–23.25), compared to mothers who perceived their milk secretion as insufficient.

These findings suggest that, beyond awareness alone, maternal perception and confidence regarding milk adequacy may play an important role in sustaining EBF practices. Mothers who reported adequate or profuse milk production were more likely to practice EBF than those who perceived their milk production as insufficient. These associations remained significant after adjustment for potential confounders (Table 4). These findings may indicate the importance of addressing maternal perceptions of milk sufficiency through breastfeeding counselling and lactation support as part of strategies to promote and sustain EBF.

## Discussion

The present study examined infant feeding practices and factors associated with early initiation of breastfeeding (EIBF) and exclusive breastfeeding (EBF) among Bhumij mothers. According to the National Family Health Survey-4 (NFHS-4), 45.3% of mothers from Scheduled Tribe communities in West Bengal initiated breastfeeding within one hour of birth [28–31], whereas 72.9% of mothers in the present study reported breastfeeding initiation within the first hour after delivery. EBF during the first six months was practiced by 65% of Bhumij mothers, exceeding the South Asian estimate (60%; UNICEF, 2024), the national estimate (64%; NFHS-5), and the state-level prevalence for West Bengal (52.3%). Compared with regional and global reports, the present findings indicate relatively higher rates of early initiation and EBF [32–37].

Although 76.5% of mothers demonstrated adequate knowledge of exclusive breastfeeding (EBF), 65.16% reported practicing EBF for the recommended first six months. This difference suggests that factors beyond breastfeeding knowledge may influence EBF practices. Previous studies have similarly reported associations between EBF and maternal confidence, family support, cultural norms, and access to breastfeeding support services [38]. The findings of the present study support the importance of considering these broader influences when designing interventions to promote and sustain EBF.

The findings of this study indicate that mode of delivery and immediate postnatal feeding practices were associated with early initiation of breastfeeding. Consistent with previous studies, caesarean delivery was associated with lower odds of initiating breastfeeding within the first hour after birth [21,39,40]. These findings highlight the potential value of targeted breastfeeding support during the immediate postoperative period to facilitate recommended newborn feeding practices [22,24,41]. The observed association between infant sex and breastfeeding initiation is also consistent with findings from some previous studies [42,43], suggesting that infant feeding practices may vary across sociocultural contexts. Taken together, these findings underscore the importance of considering delivery-related and early postnatal factors when promoting timely breastfeeding initiation. Strengthening breastfeeding support during the postnatal period, particularly following caesarean delivery, may help facilitate recommended breastfeeding practices within the study population.

In this analysis, delayed initiation of breastfeeding was observed alongside prelacteal feeding practices and colostrum discard, patterns that have also been reported in previous studies across diverse settings. These findings indicate that cultural practices and community traditions are important components of early neonatal feeding behaviours among ethnic populations in India [20,36,44–46].

Maternal knowledge of exclusive breastfeeding was strongly associated with EBF practices. Mothers who were aware of the recommended duration and benefits of EBF were more than six times as likely to breastfeed their infants exclusively (AOR = 6.14, 95% CI: 2.80-13.46). This strong association is consistent with earlier studies from diverse settings, which identify maternal knowledge as an important determinant of breastfeeding practices [47–48]. Studies from Ethiopia, Uganda, and South Asia similarly report that mothers who are well informed about EBF are more likely to practice recommended infant feeding behaviours, independent of socio-demographic and healthcare-related factors [49]. Comparable findings from systematic analyses suggest that improved breastfeeding knowledge may enhance maternal self-efficacy, facilitate early problem-solving, and strengthen motivation to continue EBF [50–52].

The current study also highlights self-perception of breastmilk secretion as an important determinant of EBF practices. Mothers who perceived themselves as having adequate milk secretion had markedly higher odds of practicing EBF (AOR = 6.12, 95% CI: 3.00-12.48). This association was even stronger among mothers who perceived profuse milk secretion, who demonstrated substantially increased odds of maintaining EBF (AOR = 7.71, 95% CI: 2.56–23.25). Maternal confidence in breastmilk production also showed a strong association with EBF [53,54].

This finding is consistent with studies from low- and middle-income countries indicating that perceived insufficient milk is among the most commonly reported reasons for early discontinuation of EBF [55]. Perceptions of milk insufficiency may be shaped by a range of factors, including early breastfeeding experiences, access to breastfeeding support, and prevailing cultural and caregiving practices related to infant feeding. The current findings therefore highlight the potential value of early lactation counseling to help mothers better understand newborn feeding patterns, manage breastfeeding-related concerns such as engorgement or latch difficulties, and receive reassurance regarding perceived milk adequacy.

## Conclusion

This study provides novel evidence on early initiation of breastfeeding and exclusive breastfeeding practices among Bhumij mothers in eastern India, a population that has been underrepresented in breastfeeding research. The findings demonstrate relatively high rates of early breastfeeding initiation and exclusive breastfeeding compared with regional and state-level estimates, while also highlighting the important role of perinatal experiences, maternal knowledge, breastfeeding confidence, and community caregiving practices in shaping infant feeding behaviours.

These findings suggest that strategies to improve breastfeeding practices in similar settings may benefit from combining breastfeeding education with timely postnatal support and interventions that address maternal concerns related to breastfeeding and milk adequacy. These findings also suggest that knowledge-based counselling may be more effective when combined with practical postnatal breastfeeding support and family-inclusive approaches during the immediate postpartum period. Further research is needed to explore the contextual and sociocultural factors influencing breastfeeding behaviours in indigenous communities.

Taken together, these findings provide context-specific evidence on breastfeeding practices among Bhumij mothers and highlight the influence of healthcare-related, maternal, and caregiving factors on early initiation and exclusive breastfeeding. These may further help inform the nation-wide maternal and child health programmes and breastfeeding promotion strategies in tribal and rural populations across India. By documenting the interaction between healthcare access, maternal perceptions, and community caregiving practices, the study offers insights that may support the development of more inclusive and culturally responsive national breastfeeding policies and interventions tailored to the needs of indigenous communities. Overall, the study contributes important insights into the social, cultural, and healthcare-related dimensions of breastfeeding practices among Bhumij mothers and underscores the need for context-specific maternal and child health strategies that build upon existing community strengths and caregiving traditions.

## Data Availability

Data is already added in the form of tables. Raw Data will be made available to academic researchers following a reasonable request.

## Acknowledgements

The authors would like to extend deepest gratitude to all the mothers for participating in this study and voluntarily contributing their time and insights. We also thank medical officers, the ASHA workers and ANMs of Primary Health Centres of Baghmundi block, Purulia, West Bengal, India for their continuous assistance during data collection.

## Author contributions

DD: conceptualisation, worked on the study design, data collection, data analysis, data interpretation, and writing of the original draft. CBM: Data interpretation, supervised and contributed to manuscript writing, review and editing. BPS: Data analysis, review and editing. ARB: conceptualisation, worked on the study design, data collection, data analysis, data interpretation, supervision, review and editing. All authors read, reviewed and approved the final version of the manuscript.

## Data availability

Data will be made available to academic researchers following a reasonable request.

## Ethical approval

Ethical approval for this study was obtained from the Institutional Ethical Committee for Biomedical and Health Research involving Human Participants, University of Calcutta (Ethical Clearance No. CUIEC/02/16/2022-23, Dated January 5, 2023). Written and verbal both informed consents were obtained from all participants prior to data collection for the study. For pre-literate respondents, consent was recorded via left thumb impression in the presence of an independent witness. The study followed the principles of the Declaration of Helsinki of 1964, as revised in 2000.

